# FREQUENCY OF ELEVATED BLOOD PRESSURE ASSOCIATED WITH LIFESTYLE CHOICES AMONG SCHOOL GOING ADOLESCENTS IN KARACHI: A CROSS SECTIONAL SURVEY

**DOI:** 10.1101/2020.05.11.20098822

**Authors:** Felicianus Anthony Pereira, Jumana Abdul Qadir, Sajida Kamran PT

## Abstract

**Background:** Studies have suggested that atherosclerotic changes take place in the body since childhood due to altered dietary patterns and sedentary lifestyle. It is critical to identify gap areas and update current literature to produce effective changes in our lifestyle.

**Study design:** A cross-sectional survey conducted among school going adolescents in Karachi.

**Methods:** A cross sectional study was performed in three different schools of Karachi. A sample size of 288 was drawn through non-probability, purposive sampling technique. Students were given a questionnaire comprising of questions regarding their physical activity levels, dietary patterns and knowledge regarding blood pressure. Blood pressure and Body Mass Index data was also recorded.

**Results:** Mean age of participants was 14.10 ± 1.097. Of the 288 students that participated in this study (122 boys and 166 girls), 227 were normal for hypertension status (93 boys and 134 girls), 27 were pre-hypertensive (7 boys and 20 girls), and 34 were hypertensive (22 boys and 12 girls). Mean systolic blood pressure was 112.73 ± 13.49, and mean diastolic blood pressure was 71.25 ± 13.03. Awareness among participants was high regarding hypertension being linked to the foods they consumed (62.8%).

**Conclusion:** Our study did not show strong correlation between physical activity and dietary patterns, with status of hypertension. Screening programs should be conducted in schools to monitor blood pressure and body mass index. High risk groups should be approached and advised for lifestyle modification.

## Introduction

With a worldwide prevalence of 40.8%, hypertension (HTN) is a rapidly increasing public health problem. It is now recognized as one of the major risk factors for cardiac, cerebrovascular and chronic kidney disease.^1^ Every year, elevated blood pressure leads to 54% of strokes, 47% of ischemic heart disease (IHD) and 7.6 million premature deaths.^2^ There is limited updated information on the prevalence of hypertension in Pakistan.^1^ In recent times, HTN has been started to be acknowledged as an important pediatric crisis.^3^ The definition of “normal blood pressure” is given as Systolic Blood Pressure (SBP) and Diastolic Blood Pressure (DBP) values <90th percentile according to age, sex and height. “Prehypertension” or elevated blood pressure for adolescents is defined as BP ≥120/80mm Hg to <95th percentile, or ≥90th and <95th percentile, whichever was lower.^4^ Evidence documents that lifestyle choices in childhood associated with risk factors which include dietary habits and physical activity levels contribute to elevated blood pressure in children.^5^ Furthermore, early origin of risk factors during adolescence and death due to IHD is also increasing. However, knowledge regarding this growing epidemic and its high prevalence is deficient among adolescents.^2^

A survey conducted by the World Health Organization (WHO) in 2010 revealed that 81% of youths between ages of 11-17 were not sufficiently physically active. Girls performed fewer physical activities than boys (84% vs. 78%). WHO claims that sedentary behavior at home contributes to the low levels of physical activity. Similarly, rise in the use of passive modes of transportation is also responsible for insufficient physical activity.^6^

Dietary intake characteristics like; frequency of eating, portion sizes, energy density, high fat, sodas and sugar intake contribute to altered dietary patterns.^7^ Pathological studies show that there is a positive correlation between atherosclerotic lesions and risk factors such as low density lipoprotein cholesterol, triglyceride, SBP & DBP, body mass index (BMI), and cigarette smoking.^5^ Hence, it is important to evaluate diet for these characteristics in adolescence because it predicts dietary intake in adult life which can thereby cause a risk of chronic diseases.^7^

The purpose of this research was to assess the frequency of elevated blood pressure associated with lifestyle choice in school going adolescents and assess awareness of the modifiable risk factors.

With rate of mortality due to non-communicable diseases rising to 52 million by 2030 it is crucial for our present generation of adolescents to be aware of elevated blood pressure and their relative lifestyle.^2^

According to author’s knowledge, elevated blood pressure in adolescents is under reported in our part of the world.^8^ Two major contributing factors are: altered dietary patterns due to ease of access to fast food,^9^ and increased screen time leading to sedentary lifestyle.^7^

## Methods

Permission to conduct this study was obtained from Institute of Physical Medicine & Rehabilitation, Dow University of Health Sciences. (Reference no. IPM&R/DUHS-17/061)

The cross-sectional survey was conducted on children aged 13 to 16 and enrolled in public and private schools in Karachi, Pakistan. It was performed in compliance with declaration of Helsinki. Principals, teachers and students were briefed about the study. They were explained about the purpose of the study and were assured strict confidentiality. Consent was obtained from parents, as participants in the study were below legal age. Children, teachers, and parents (through consent forms) were informed of the procedure of the study. Sample size was calculated at 282 with confidence interval of 95% using OpenEpi version 3.01. Data was obtained via a self-design questionnaire.

Blood pressure was measured using a mercury sphygmomanometer (Certeza Desk Type CR-2001). Children were in a calm environment before readings were taken. Appropriate cuff size was used during the research. Blood pressure measurements were taken with the arm extended over the table, at the level of the heart. ^10^

Two readings were taken with 5 minutes’ interval between each reading, and the average was calculated. ^10^ Classification of BP percentiles was determined using normative tables obtained from National Health and Nutrition Examination Survey data. The 95th percentile was used as a measure of elevated blood pressure for the adolescents’ gender, height and weight.

Weight was measured using an electronic weighing scale (Senior Model DB-6020 - accurate to 0.1kg). Children were weighed wearing light clothes and no shoes. Standing height was measured with shoes removed and the child facing away from the wall, with the heels, buttocks, shoulders and head touching the wall and the child looking ahead and the external auditory meatus and lower margin of the orbit aligned horizontally. ^2^ Height was measured using a tape measure (accurate to ±1/2cm). Measurements were recorded between 8:30 a.m. and 11:00 a.m. ^11^ Weight and height were converted to metric measurements to determine the BMI, which is calculated by weight (kg) divided by height squared (m^2^).

## Results

Children included in the study were between 13 to 16 years of age. Mean age of participants was 14.10 ± 1.097. Of the 288 students that participated in this study, 27 were pre-hypertensive, and 34 were hypertensive (9.4% and 11.8% respectively). (Table 1) Gender wise distribution of status of HTN is shown in Figure 1. Mean SBP was 112.73 ± 13.49, and mean DBP was 71.25 ± 13.03.

**Table 1:** status of HTN.

|  | Frequency | Percent |
| --- | --- | --- |
| Status of HTN |  |  |
| Normal | 227 | 78.8 |
| Pre-hypertensive | 27 | 9.4 |
| Hypertensive | 34 | 11.8 |
| Total | 288 | 100 |

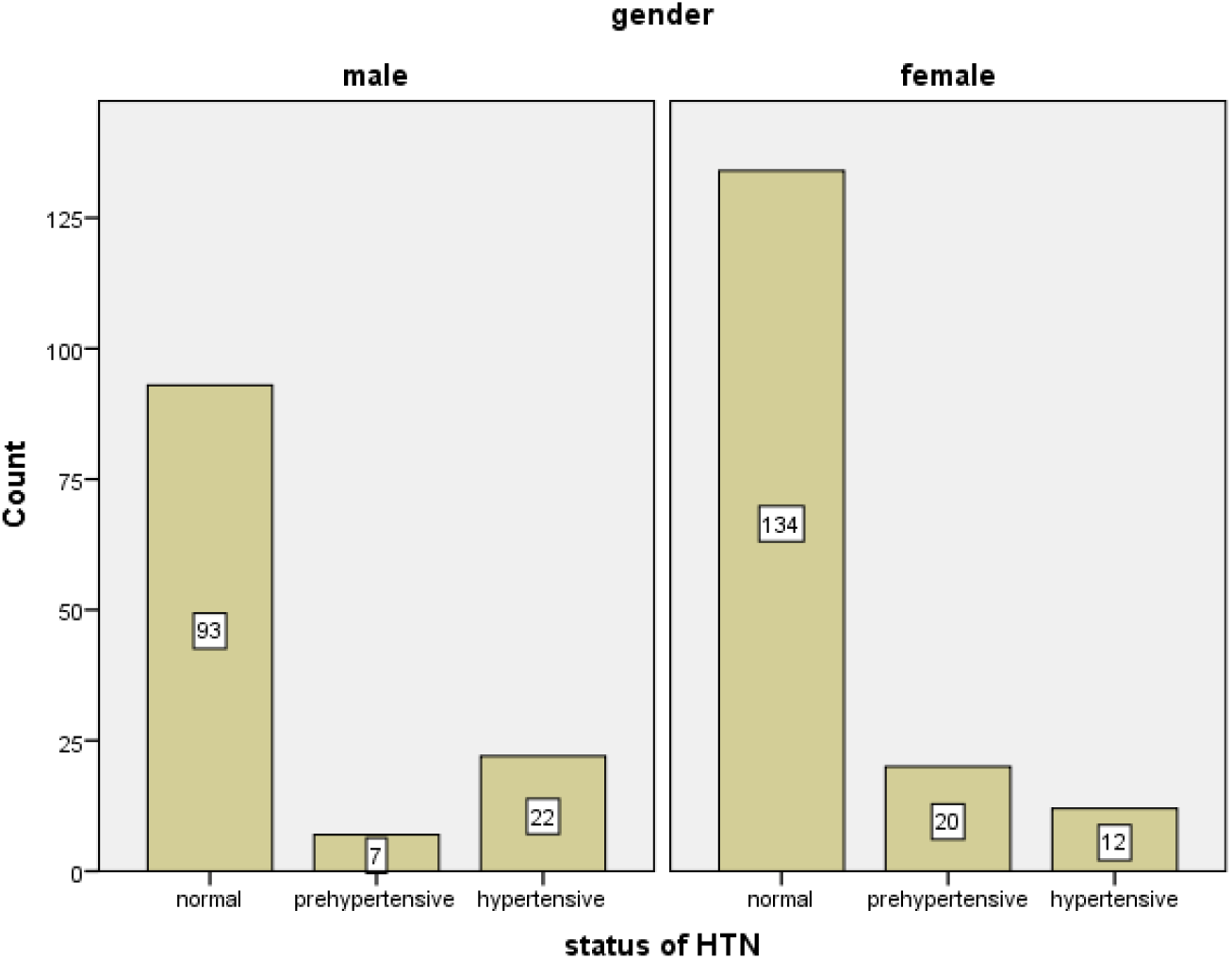

The association found between status of HTN and status of BMI was statistically significant (p value=0.000). (Table 2) Cross tabulation of status of hypertension with frequency of sitting and lying down showed no statistical significance (p-value=0.878). Statistical significance (p-value=0.001) was also noted between cell phone usage and status of hypertension. Weak association (p-value=0.936) between cigarette smoking and status of hypertension was noted in this study. 285 participants did not smoke at all. 2 participants smoked to a moderate extent; both had normal HTN status. Statistical significance (p-value=0.007) was found between consummation of soft drinks, and the status of hypertension (Table 3)

**Table 2:**
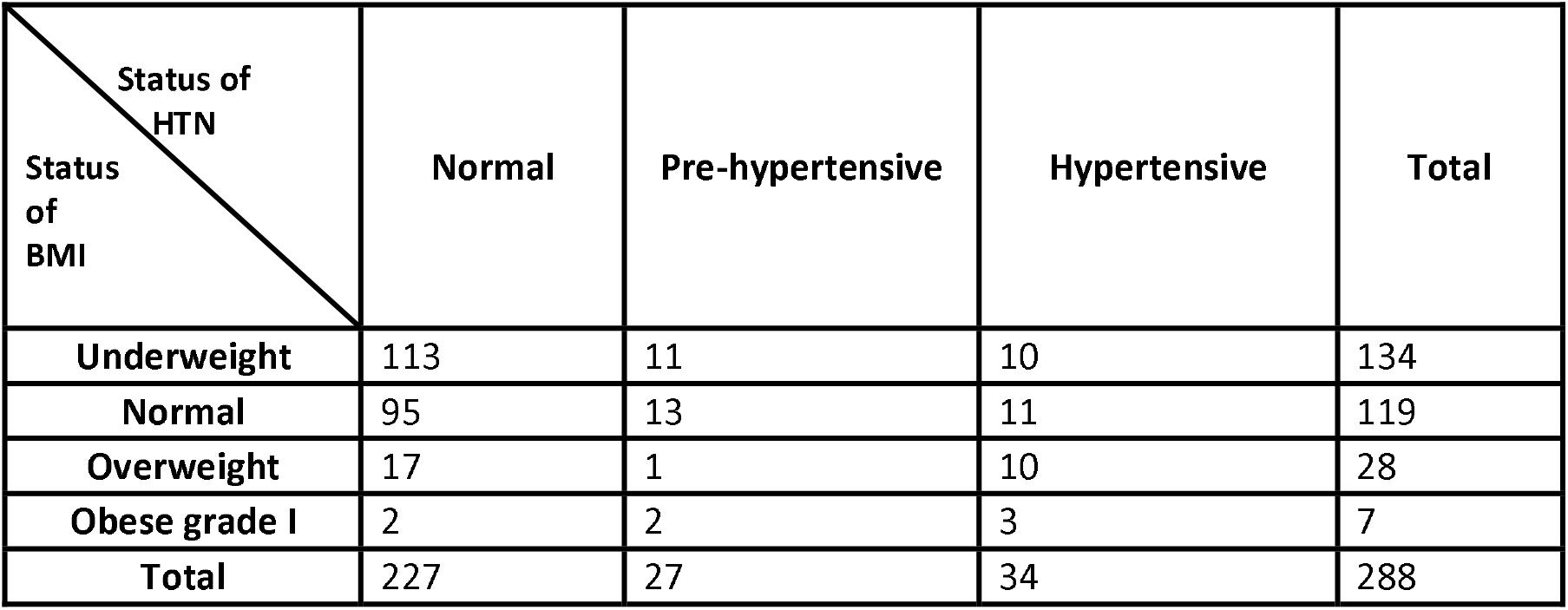
association of status of BMI with status of HTN.

| Status of HTN<br>Status of BMI | Normal | Pre-hypertensive | Hypertensive | Total |
| --- | --- | --- | --- | --- |
| Underweight | 113 | 11 | 10 | 134 |
| Normal | 95 | 13 | 11 | 119 |
| Overweight | 17 | 1 | 10 | 28 |
| Obese grade I | 2 | 2 | 3 | 7 |
| Total | 227 | 27 | 34 | 288 |

**Table 3:**
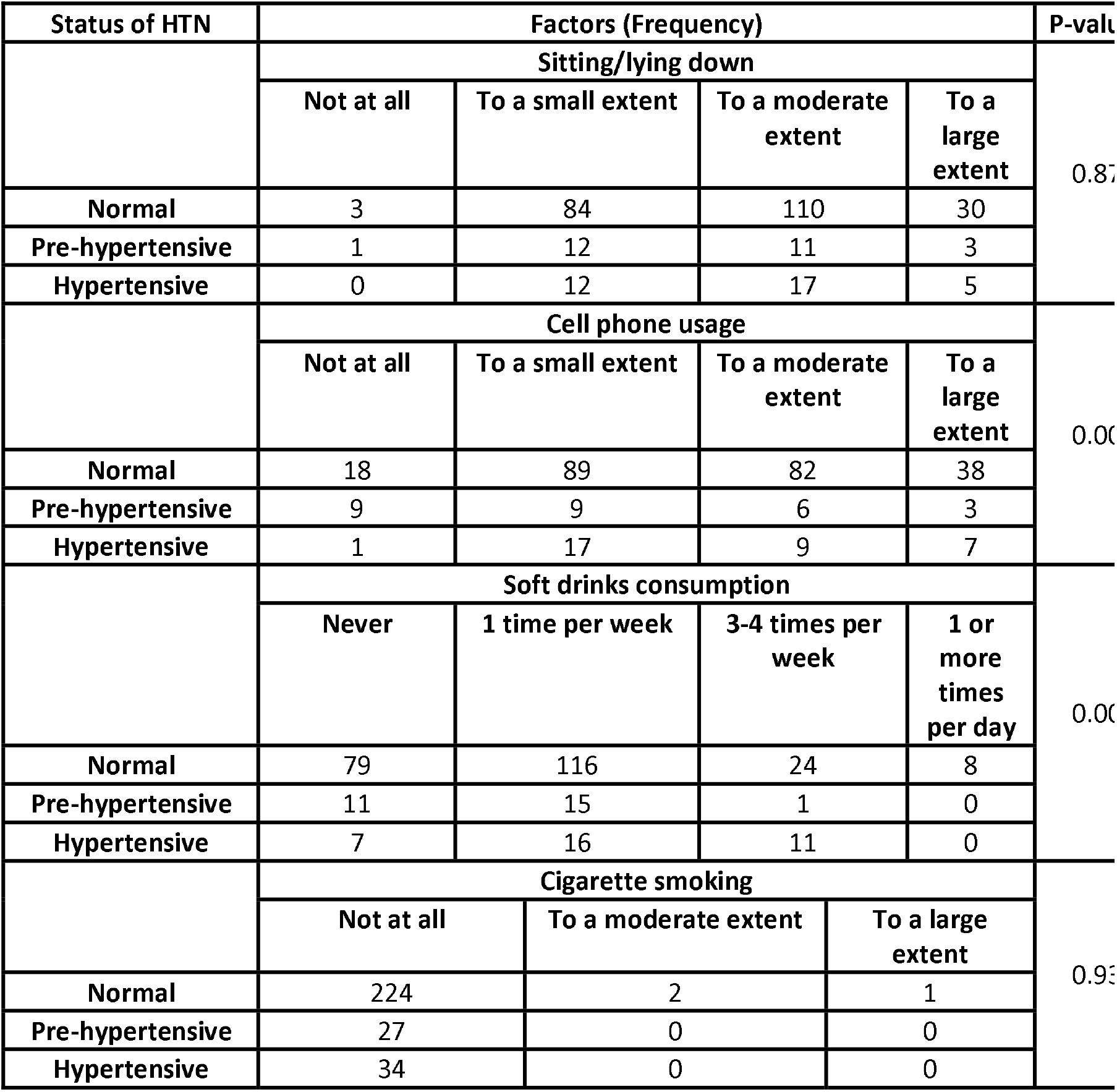
association of status of hypertension with sitting/lying down, cell phone usage, soft drinks consumption and cigarette smoking.

## Discussion

In this study the prevalence of BP > 95th percentile is 11.81%, BP between 90th and 95th percentile is 9.38%. These are quite similar with the results of previous studies.^12 13^ Various studies in different parts of the world report a prevalence ranging from 0.46% to 21.8%. The reason for this might be due to the difference in the study group’s age and ethnicity.^14^ It is worth mentioning that the difference in number of times the measurements were taken reduced the prevalence from 13% to 1% between the first and third visits.^15^

In this study, obesity, altered food habits and physical inactivity are three risk factors that have been found to be independently associated with high BP. Results of a study conducted by Cinteza and Balgradean and Rahman et al. support the association between obesity and elevated blood pressure.^16^ A study conducted on Pakistani students had the same findings.^17^ Another study suggested that there is a shift in the arterial pressure control mechanism of diuresis and naturesis due to activation of sympathetic nervous system, in obese adolescents which ultimately leads to higher BP levels.^18^ In our study no statistically significant association was noted between consumption of junk food and that status of HTN. Similar results were found in a study conducted by Jasmine s sunder et al.^19^ However, in a study conducted by Sentamizh Prasad et al. there was a significant statistical association between the former two variables.^2^ As with the results of other studies, our research indicated strong statistical significance between increased screen time and elevated blood pressure.^20 21 18 22 23 24^ Due to reduced physical activity, there is dysregulation of body weight, insulin usage and indirectly blood lipids, glucose and clotting factors get deranged, altering the blood vessels and thereby altering blood pressure.^25^

Knowledge about HTN was high among participants. 89.2% had been informed by a teacher or a guardian about BP; this contrasts with the findings of Prasad et al study, where adolescents had a poor understanding of knowledge of BP.^2^ Majority of participants (62.8%) were aware of BP being linked to the foods they consumed.

Limitations of our research were that BP measurements were taken twice; assessment of dietary patterns was done via questionnaire, based on responses from participant’s memory, which may be subject to recall bias. No observation was done to assess levels of physical activity. Also, a stratified sampling technique would have been more appropriate, as the participants in our study were from Dawoodi Bohra community, and is not a complete representation of the different ethnic groups of adolescents in the country.

Informative sessions can be conducted in schools to inform children and parents about relation between obesity and hypertension. Intervention strategies should be implemented at national level, such as the banning of soft drinks at schools by the Punjab government^1^ to reduce the chances of cardiovascular diseases developing later in life.

## Conclusion

We conclude that there is no strong correlation between physical activity and dietary patterns with status of hypertension among adolescents. However, screening programs should be conducted in schools to monitor blood pressure and BMI. Ultimately, high risk groups should be approached and advised for lifestyle modification.

## Data Availability

Data regarding information of students is available with authors.

## Acknowledgement

The authors would like to acknowledge all school children and their caregivers for their co-operation in the study.

## Conflict of interest

The authors report no conflict of interest.

## Funding

No funding was obtained for this study.

## Notes

### Competing Interest Statement

The authors have declared no competing interest.

